# Intracerebral Hemorrhage etiological classification systems and their correlation with neurological deterioration

**DOI:** 10.1101/2023.08.14.23294102

**Authors:** Daniel Vázquez-Justes, Gerard Mauri-Capdevila, Gallego Yhovany, Miriam Paul-Arias, Raquel Mitjana, Francisco Purroy

## Abstract

**Introduction:** Unlike ischemic stroke, the etiological classification of patients with intracerebral hemorrhage (CH) lacks consensus. Our study focuses on two commonly-used classification systems: SMASH-U and H-ATOMIC. The main difference between them lies in the fact that the H-ATOMIC system considers the simultaneous presence of different etiologies in a single patient. The association between the two classifications with relation to neurological deterioration (ND) and clinical outcomes remains so far unexplored.

**Methods:** We recruited consecutive ICH patients from 2015 to 2022, determining etiology was on discharge. Demographic, radiological and clinical characteristics were recorded. ND during hospitalization in the 7 days after stroke was the main clinical endpoint.

**Results:** 301 patients were recruited. 124 patients (41.2%) experienced ND. The hypertensive subtype was the most frequent etiology with both classifications. In 149 (49.5%) more than one possible etiology for the ICH were recognized. The most frequent combination was hypertension + either probable or possible amyloid angiopathy, in 64 patients (21.3%). Significant differences in ND proportions were observed across groups with both systems. ICH related to anticoagulation was associated with a greater risk of ND: 63.5 % in SMASH-U and 62.5% in patients with a combination of Hypertension and Oral Anticoagulants in H-ATOMIC. Both these etiological groups and that containing combined etiologies were statistically significant according to multivariate analysis. Intraventricular extension, blood pressure control and initial volume were also related to ND.

**Conclusion:** Etiology of the ICH could be related to the risk of ND during hospitalization. Anticoagulation-related etiologies may present the highest risk, especially when combined with hypertension.

## Introduction

Spontaneous Intracerebral Hemorrhage (ICH) is present in approximately one in ten patients with stroke [1, 2]. It is associated with a significantly poorer prognosis and higher morbi-mortality than ischemic stroke (IS)[1-3]. Aging, male sex, hypertension, illicit drug use, oral anticoagulation treatment and alcohol consumption have all been identified as risk factors for ICH [1]. Hypertension and Cerebral Amyloid Angiopathy (CAA) are the most frequent causes of ICH [1, 2, 4]. In contrast to IS, in which etiological classifications of the type used in Trial Org 10172 in Acute Stroke Treatment [5] are widely used, there is no consensus on which etiological classification should be used in ICH patients [1]. Moreover, the most widely-used predictive models in ICH do not consider the etiology [6, 7].

Until 2012, classifications of ICHs had mainly considered their anatomical location. In 2012, SMASH-U [8] became the first system proposed for the etiological classification of ICH. It included six categories: hypertension, CAA, systemic etiology, medication, structural, and unknown. This classification allows for quick and easy application in a clinical setting. It has been related not only to 3-month mortality [8] but also to hematoma expansion [9] and in-hospital mortality [10]. This system considers one etiology to be the most likely culprit, even when more than one possible cause is present.

In 2016, the H-ATOMIC etiologic classification was proposed [11]. The categories in this classification included hypertension (H), amyloid (A), tumor (T), oral anticoagulation (O), malformation (M), infrequent (I) and cryptogenic(C). The main difference with respect to the SMASH-U classification is that it allows the inclusion of different simultaneous etiologies. A patient could be classified in different ways according to the degree of certainty: definite, probable or possible. For an etiology to be considered “definite”, the absence of any other possible etiology would be required.

The concordance between these two systems was analyzed in a previous study [12]. However, the influence of different combinations on the clinical outcome was not studied. How different causes of ICH affect outcomes remains an issue yet to be adequately explored. ICH patients are at high risk of Neurological Deterioration (ND) during the acute phase; this can be due to several different factors, including hematoma expansion or intraventricular extension [13]. ND is known to be a determinant of outcome in patients with ICH. However, the association between etiologic subtypes of ICH and ND has yet to be studied in sufficient depth. It would be interesting to investigate whether the coexistence of multiple etiologies in the same patient leads to a worse prognosis. Establishing a correlation between ICH etiologies and early prognosis could help practitioners to make individual decisions regarding patient management.

We sought to compare how ICH patients were differently classified according to the system applied and how different etiologies combined. We also wanted to study the relationship between ICH etiology, as defined by the SMASH-U and H-ATOMIC systems, and the occurrence of ND during hospitalization.

## Methods

### Data availability statement

Requests for access to the data reported in this article will be considered by the corresponding author.

### Design and study population

The study was conducted following the STROBE guidelines. We prospectively included consecutive adult patients (>18 years) with spontaneous ICH who were attended to in the emergency room of our university hospital either within 8 hours from the onset of symptoms or who had previously been seen asymptomatic between April 2015 and March 2021. The local ethics committee approved the study (HUAV, code: 2168), and written informed consent was obtained from all participants. ICH was diagnosed according to an initial CT scan. Patients with stroke but without intracranial bleeding were excluded from the study, as were those exhibiting a hemorrhagic transformation from an ischemic stroke, even though previous thrombolysis had been administered. Patients with intracranial non-parenchymal bleeding (subdural, epidural and subarachnoid hemorrhage) and those whose ICH was secondary to head trauma were also excluded. All patients underwent a CT scan during the initial evaluation in the emergency department. Other neuroimaging techniques, such as CT angiogram, MRI, MRI angiogram and digital subtraction angiography, were performed at the discretion of the stroke neurologist. Information relating to how the etiological work-up was conducted is presented in the figure.

The location of the ICH according to basal CT scan were: deep location, if the bleeding was in the thalamus, internal capsule or basal ganglia; lobar or cortico-subcortical; infratentorial for both brainstem and/or cerebellar locations; pure intraventricular location; simultaneous ICH, if there was more than one ICH on the initial CT scan; or mixed location, when parenchymal ICH had intraventricular extension. Hematoma volume was measured using the standard ABC/2 method [14].

Information about demographic characteristics, previous medication, laboratory data, and stroke severity (Glasgow Coma Scale and NIHSS scale) were recorded. Outcome at three months was evaluated by the attending physician and recorded on a medical chart. The admission destination of the patient: stroke unit, Intensive Care Unit (ICU), or conventional hospitalization was documented. Information about medical and surgical management in the acute phase and during hospitalization was also recorded. The blood pressure (BP) measurement upon admission to the emergency department was also registered. Optimal BP control was considered in those patients who were hemodynamically stable and whose systolic BP (SBP) was below 140mmHg during the first six hours after admission, regardless of the number and type of medications required to reduce their BP.

Hematoma expansion was defined as an absolute of more than 6 mL, or a relative increase of more than 33% with respect to their baseline CT [15].

### Classification of the ICH etiologies

The SMASH-U [8] classification includes the following groups: structural vascular lesions (S), medication (M), Cerebral Amyloid Angiopathy (CAA), systemic disease (Sys), hypertension (H), and unknown (U). On the other hand, the H-ATOMIC [11] system criteria include: hypertension (H), Cerebral Amyloid Angiopathy (CAA), tumor (T), oral anticoagulants (O), malformation (M), infrequent (I), and cryptogenic (C). Etiology subtypes were determined by the attending physician at discharge. A vascular neurologist (DVJ) subsequently reviewed the cases and defined the different subtypes according to both the aforementioned classifications. As in a previous study [12], given the large number of possible combinations in the H-ATOMIC system, we applied a simplified approach.

In the SMASH-U system, the hypertensive subtype was considered in all patients with hypertension and deep location, or hypertension and other etiologies discarded. In the H-ATOMIC system, definite hypertensive (H1) was only considered in patients without any other possible concomitant etiology apart from hypertension.

Probable CAA (A2) was diagnosed when the patient met the corresponding Boston criteria. Possible CAA (A3) was considered when possible Boston criteria were present [11]. In contrast, lobar ICH in patients over 55 years old accompanied by deep microbleeds on MRI was considered to constitute a combined etiology of hypertension and CAA (H2A3 or H3A3 according to H-ATOMIC system). In the cases in which an MRI was not available, lobar ICH was classified as possible CAA (A3) according to the H-ATOMIC system, but as CAA using the SMASH-U system, if the patient was over 55 years old and there was no evidence of arteriovenous malformation.

When an arteriovenous malformation was detected in the study, structural subtype (SMASH-U), or definitive M1 (H-ATOMIC), was considered.

In the case of cerebellar ICH, CAA was considered if there were strictly lobar microbleeds associated on the MRI [16] as with A2 in H-ATOMIC and CAA in SMASH-U.

In order to elucidate whether the combination of microangiopathy and the use of anticoagulants had any impact on the clinical outcome, we decided to create two combination groups in the H-ATOMIC system: one relating to anticoagulants and hypertension (HO); and another for anticoagulants and possible or probable CAA (AO). The HO subtype therefore encompassed various combinations of hypertension and the use of anticoagulants: the H2O2, H3O2, H2O3 and H3O3 groups of the H-ATOMIC [11], and the AO subtype included: A2O2, A3O2, A2O3 and A3O3. We also defined a group that combined patients who had possible or probable CAA and hypertensive microangiopathy (HA). This latter group encompassed the H2A2, H3A2, H2A3 and H3A3 subgroups defined in the original H-ATOMIC classification.

Finally, any other combination of etiologies, apart from those already mentioned, was included in a group that we called “combined”. For the statistical analysis, we analyzed these different combinations as independent groups. The composition of these groups is provided in the Supplementary Material.

### Outcomes

Patients were subjected to systematic follow-ups at 24 hours, seven days, discharge, and 90 days. The primary outcome was ND which was defined as an increase of 4 or more points in the *National institute of Health Stroke Scale* (NIHSS) score from baseline to 7 days or as a decrease of one or more points on the *Glasgow Coma Scale* (GCS) registered during the first 7 days after the onset of symptoms, as in previous studies [17]. As a secondary outcome, we recorded major disability at three months, which was defined as a score of 3 to 6 on the modified Rankin scale.

### Statistical analysis

We determined differences in the distribution of each etiologic subtype in both classification systems. Demographic characteristics, vascular risk factors, stroke severity, clinical evolution, management, functional outcome and ND were all analyzed for each etiological group using both classification systems. For this purpose, continuous variables were compared between different groups, using either an ANOVA test or the Mann-Whitney U test. The qualitative variables were then compared using the chi-squared test or Fisher’s exact test when the expected cell frequency was <5.

We used multivariable logistic backwards stepwise regression analysis to obtain adjusted odds ratios with which to assess the contribution of the different variables to ND. We included the variables for which p<0.05 in univariate testing. Logistic regression was conducted twice: once with SMASH-U and then repeated with H-ATOMIC etiologies. The statistical analysis of the data was carried out using version 20.0 of the SPSS statistical package (SPSS, Chicago, IL, USA).

## Results

301 patients with ICH were included in the study. The mean age was 71 (SD 13.2) years old, 118 (39.2%) were female, 124 patients (41.2%) suffered ND, and 115 (38.2%) died within three months. For one patient, the clinical outcome was not known, due to loss of follow-up.

The differences in classification between systems are presented in Table 1. With both classifications, the most frequent etiology was hypertensive: 135 patients (44.9%) were classified as hypertensive using SMASH-U. In 100 of these patients (74.1%), no other etiology was found apart from hypertension and these patients were classified as H1 using H-ATOMIC. 68 patients (22.6%) were classified as CAA using SMASH-U. Of these, only 13 patients (19.1%) met the Boston criteria for probable CAA and could not be associated with any other possible etiology (A2); 42 (61.8%) had additionally hypertension as a possible cause (HA); and in 3 patients (4.4%), there was evidence of additional previous anticoagulant use (A2O2 or A2O3) and so they were classified as AO.

15 patients were classified as exhibiting a systemic etiology according to SMASH-U. 7 (46.7%) of these 15 patients exhibited an infrequent etiology according to H-ATOMIC, while the other 8 (53.3%) were included in the combined group.

16 (5.3%) patients remained with an undetermined etiology according to SMASH-U, whereas 7 (2.3%) were cryptogenic according to H-ATOMIC.

The observed differences in the proportions of the demographic characteristics, previous treatments, stroke severity, BP management, neuroimaging and outcomes across the different etiological subtypes observed using the two classification systems have been summarized and are presented in Table 2 and Table 3.

Stroke severity as assessed by the NIHSS scale and GCS score did not differ significantly between groups. Systolic BP levels on admission were higher amongst the hypertensive, CAA and medication groups in SMASH-U (p<0.001), and in H1 and HA in H-ATOMIC (p<0.001). Volume was higher in the CAA group in SMASH-U (median of 31.5 ml), with the greatest volume being registered in the AO group of H-ATOMIC (median of 64ml). Hematoma expansion was more frequent in the medication group in SMASH-U (25.4%) and in the HO in H-ATOMIC (31.4%). For 60 patients, there was no CT scan at 24 hours with which to evaluate hematoma expansion.

ND was most frequent in the systemic and medication groups in SMASH-U, whereas in H-ATOMIC, it was most frequent in O1 and HO. Similarly, the functional outcome at three months was poorest in the medication group in SMASH-U, and also in O1 and HO groups in H-ATOMIC.

The factors that were related to ND are presented in Table 4. The patients with ND were older (p<0.001), had more severe strokes (higher NIHSS score), had higher volumes of ICH on the initial neuroimaging (p<0.001), had longer hospitalizations (not shown in the table, p<0.001), had less frequently optimally controlled BP, and more frequently showed intraventricular extension (p<0.001). After multivariate analysis (Table 5), volume and intraventricular extension remained related to ND after the adjustment. Optimal BP control was inversely associated with ND Regarding the etiological groups, the medication group with SMASH-U remained related to ND, whereas the HO group and combined group remained related to the H-ATOMIC system.

## Discussion

Our study highlights the difficulties involved when categorizing ICH patients etiologically, highlighting significant discrepancies between some of the subgroups in the SMASH-U and H-ATOMIC systems. About half of our sample had more than one etiology that could have contributed to the ICH, as can be observed in the distribution of etiologies according to the H-ATOMIC system. Microangiopathy, whether hypertensive or CAA, or a combination of the two, was considered responsible for three out of every four cases. As expected, the most frequent combination, which was observed in one out of every five patients, was between possible or probable CAA and hypertension. It is interesting to note that in almost two out of every three patients diagnosed with CAA using the SMASH-U criteria, there was a possible contribution of hypertension.

It has been previously reported that BP levels in the emergency room tend to significantly vary across different etiological groups [18]. However, in our sample, although the basal levels of BP varied, they did not appear to constitute a reliable factor for disclosing the underlying etiology of the ICH. In our case, patients classified associated with other etiologies rather than hypertension exhibited relatively high levels on BP on admission. Only patients belonging to the A2 and C1 groups had mean systolic blood pressure (SBP) levels that were below 140 mmHg upon admission.

We found a high proportion of patients with ND (up to 41.2%) during the first week. This rate was similar to that previously reported [13]. This was probably because we included both patients with early ND, which is traditionally defined as occurring within 24 hours of stroke onset, and delayed ND, which we considered within the first week, following previous definition [17]. The factors related to ND in our sample were similar to those reported in previous studies [13, 15, 17, 19] and included basal volume, stroke severity, and intraventricular extension. Interestingly, optimal BP control was inversely related to ND, with this relationship persisting even after conducting a multivariate analysis. BP levels had previously been related to ND [17], but whether controlling this factor has a positive impact on ND remains unknown and underexplored. This could be related to the improved outcome observed in patients undergoing intensive BP control which were described in clinical trials Interact 2 and Interact 3 [20, 21].

We found it interesting that several etiological groups remained significantly related to ND even after multivariate analysis, as occurred with both classification systems. In accordance with previous studies [8], we observed how hypertension and structural-related ICH, as well as its cryptogenic and unknown subtypes, tended to have better clinical outcomes, our results suggest this may be associated with lower rates of ND in these groups. Conversely, CAA, anticoagulation related ICH, and infrequent and systemic etiologies were associated with poor outcomes.

Among the groups based on the different combinations identified by the H-ATOMIC system, only HO remained significantly related to ND. Moreover, according to SMASH-U, the medication group was also significantly associated with poor prognosis. This observation was consistent with previous studies that had reported a relationship between anticoagulants intake and both hematoma expansion [9] and poor outcome at three months [8].

Whether anticoagulation intake should, in itself, be considered an independent etiology of ICH is a rather controversial question. It has been suggested that anticoagulation alone should not be regarded as a cause of ICH. According to this view, in cases of anticoagulation-related ICH, the presence of small vessel disease should be considered the primary contributor to ICH, with anticoagulation only being considered a risk factor for its development [22]. Our results suggest that considering a group of anticoagulant-related ICH is useful from a clinical point of view. Such patients have poorer clinical outcomes compared to those who do not take anticoagulants, even when other etiologies coexist. It is important to note that taking anticoagulants was only found to constitute an isolated etiology for ICH in 2.3% of the sample, and that the majority of patients on anticoagulation had other possible etiologies for ICH. Anticoagulation should be considered another contributing factor to ND along with other factors such as hematoma expansion, as it has been recognized [23]. Our results suggest that this is particularly relevant when anticoagulation is associated with ICH involving a hypertensive etiology. Anticoagulation has also been included in some models proposed for the prediction of ND [24]. However, the interaction between anticoagulation and microangiopathy had not previously been related to ND. Further studies are needed to determine whether this effect of anticoagulation on ND is particularly important in patients with hypertensive ICH.

Finally, we found that the group of combined etiologies was also related to ND. However, this group was too heterogeneous to allow us to draw conclusions about any specific combination of etiologies that could have significantly contributed to ND. A possible explanation is that the combination of different etiologies could, in itself, also be related to a greater risk of ND.

### Limitations

Our study has several limitations. Firstly, some etiological groups, and particularly those within the H-ATOMIC system, had relatively low sample sizes. This could have led to us missing some significant associations. In fact, certain etiological groups showed a tendency towards significance related to ND; this was the case of patients with systemic etiologies and either CAA in SMASH-U or who belonged to the HA group in H-ATOMIC. Another limitation was that not all patients underwent a brain MRI. In our study, the diagnosis of CAA was based on an early version of the modified Boston criteria [25], rather than on the more recently published Boston criteria V2.0 [26]. Moreover, the presence of hematoma expansion was unknown in many patients. We could not, therefore, accurately establish whether there were different proportions of hematoma expansion between groups, as previously reported [9].

### Conclusions

It was possible to relate the etiology to ND during the first week after ICH. The SMASH-U and H-ATOMIC etiological classification systems could be useful for identifying patients with a higher risk of ND. To be more specific, patients on anticoagulants seemed to have a particular risk of ND, especially when this was associated with hypertensive etiology. Moreover, the combination of different etiologies could, in itself, also be related to a greater risk of ND.

## Authors contributions

FP, DV conceived the study. FP, DV designed experiments. FP, DV, GM, YG, PAM cohorts’ recruitment and clinical data. RM analyzed imaging data. FP and DV participated on data interpretation and draft the manuscript. All authors critically revised the final version of the manuscript. All authors approved the final version to be published. FP procured funding.

## Declaration of interest

All authors declare no competing interests.

## Data Availability Statement

Requests for access to the data reported in this paper will be considered by the Lead contact on reasonable basis.

## Acknowledgements

We are grateful to all recruited patients, the members of Clinical Neuroscience group at IRBLleida and personal of Neurology Department at Hospital Universitari Arnau de Vilanova de Lleida for scientific discussions and instrumental help. The study was supported by a grant from INVICTUS plus Research Network (Carlos III Health Institute: RD16-0019-0017), Government of Catalonia-Agència de Gestió d’Ajuts Universitaris i de Recerca (2017SGR1628 and 2021SGR01479); Carlos III Health Institute and co-funded by European Union (ERDF/ESF, “Investing in your future” and “A way to build Europe”): PI20/01575

---

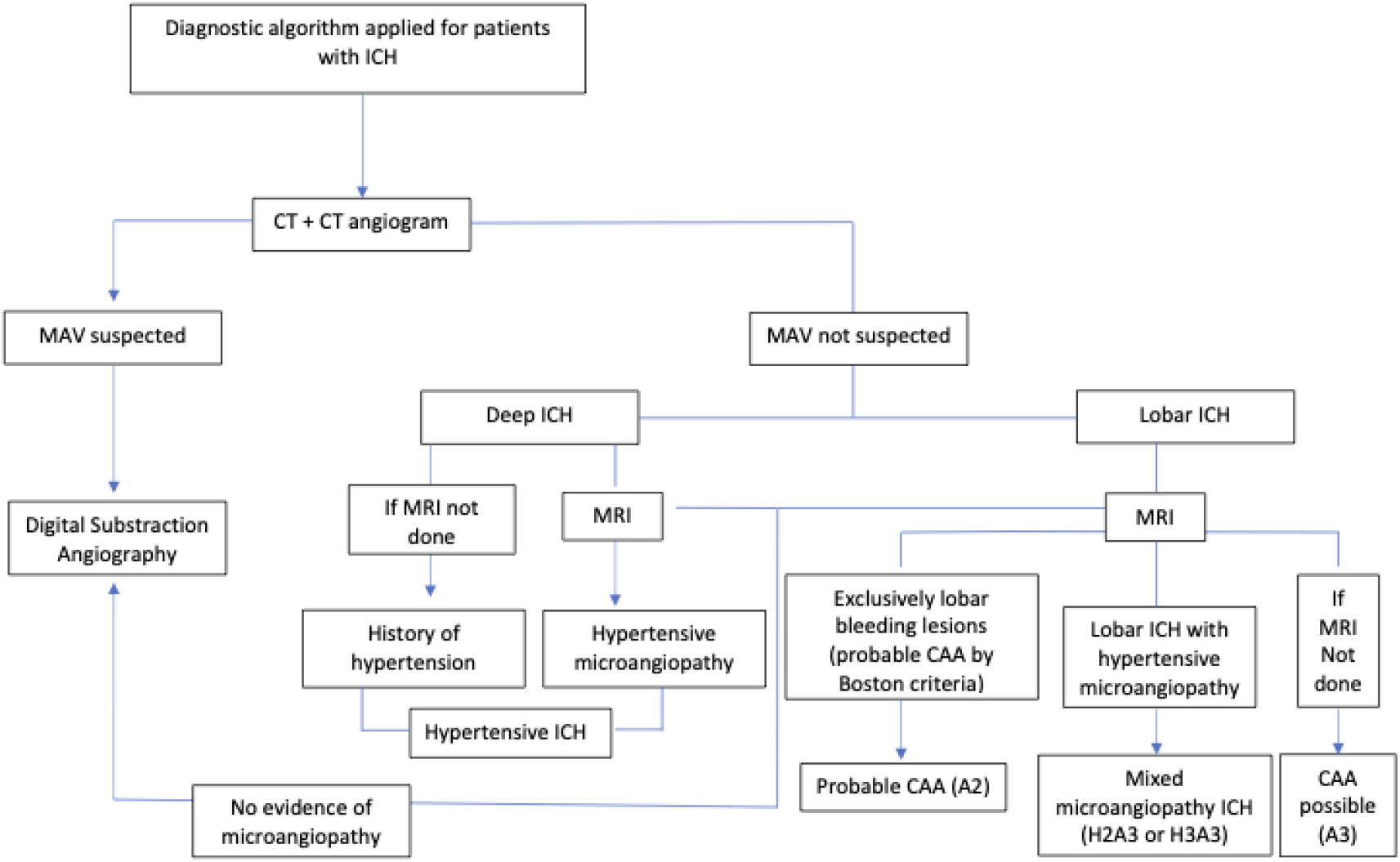

## Notes

The authors declare that they have no conflicts of interest

### Competing Interest Statement

The authors have declared no competing interest.

### Author Declarations

The ethics committee from the Hospital Universitari Arnau de Vilanova de Lleida approved the study (HUAV, code: 2168), and written informed consent was obtained from all participants.

